# Immunogenicity and safety of coadministration of COVID-19 and influenza vaccination among healthcare workers

**DOI:** 10.1101/2022.06.09.22276030

**Authors:** Isabell Wagenhäuser, Julia Reusch, Alexander Gabel, Anna Höhn, Thiên-Trí Lâm, Giovanni Almanzar, Martina Prelog, Lukas B. Krone, Anna Frey, Alexandra Schubert-Unkmeir, Lars Dölken, Stefan Frantz, Oliver Kurzai, Ulrich Vogel, Nils Petri, Manuel Krone

## Abstract

**Background:** A third dose of COVID-19 vaccination (‘COVID booster vaccination’) has become established as an important measure to strengthen the immune response against SARS-CoV-2. In contrast, seasonal influenza vaccination has been an important infection prevention measure for years, especially among highly exposed healthcare workers (HCWs). Coadministration of vaccines against COVID-19 and seasonal influenza could be an efficient strategy to protect HCWs from two major viral respiratory infections. Yet, the immunogenicity and safety of coadministration remains to be evaluated.

**Methods:** This study examines the differences in Anti-SARS-CoV-2-Spike IgG antibody formation as well as side effects based on a digital questionnaire after a third COVID-19 vaccination with or without coadministration of a seasonal quadrivalent influenza vaccine (Influvac Tetra vaccine 2021/2022). 1,231 HCWs were recruited who received a mRNA-based booster COVID-19 vaccination (mRNA-1273 or BNT162b2mRNA) after basic immunisation with BNT162b2mRNA twice. Anti-SARS-CoV-2-Spike IgG levels were determined at least 14 days after vaccination by SERION ELISA *agile* SARS-CoV-2 IgG.

**Findings:** Anti-SARS-CoV-2-Spike IgG concentrations were by 25·4% lower in individuals with coadministration of the seasonal quadrivalent influenza vaccination than without (p<0·01). There was no statistically significant difference in the reported side effects. The concentration of Anti-SARS-CoV-2-Spike IgG was higher in HCWs who had received the influenza vaccine concomitantly with mRNA-1273 than with BNT162b2mRNA as third COVID-19 vaccine (p<0·0001).

**Interpretation:** Coadministration of the seasonal quadrivalent influenza vaccine significantly limits the levels in Anti-SARS-CoV-2-Spike IgG levels, with a more restricted elevation in case of a BNT162b2mRNA booster vaccination compared with mRNA-1273 vaccine. The reduced humoral immune response in case of coadministration needs to be considered in seasonal vaccination recommendations, although the consequences of lower Anti-SARS-CoV-2-Spike IgG levels for the protection against SARS-CoV-2 infection and severe COVID-19 disease course are currently unknown. An augmented mRNA-based COVID-19 vaccine dosage may compensate for the restricted immunogenicity in case of coadministration.

**Funding:** This study was funded by the Federal Ministry for Education and Science (BMBF) through a grant provided to the University Hospital of Wuerzburg by the Network University Medicine on COVID-19 (B-FAST, grant-No 01KX2021) as well as by the Free State of Bavaria with COVID-research funds provided to the University of Wuerzburg, Germany. Nils Petri is supported by the German Research Foundation (DFG) funded scholarship UNION CVD.

**Research in context:** *Evidence before this study:* For evaluation of the previously published evidence, PubMed and medRxiv were searched for the terms “influenza vaccination”, “influenza vaccine”, “influenza”, “flu”, “seasonality”, combined with “coadministration”, “concomitant”, “COVID-19 vaccination”, “COVID-19 vaccine”, “SARS-CoV-2”, in title or abstract, published between 1^st^ of January 2020 and 18^th^ of May 2022. To date, it is unclear if coadministration of COVID-19 and influenza vaccine is effective and safe, particularly in the cohort of healthcare workers (HCWs) as key public health stakeholders. For the subunit COVID-19 vaccine NVX-CoV2373, an impairment of Anti-SARS-CoV-2-Spike IgG levels has been shown in individuals coadministered with a seasonal influenza vaccine. The two previously published studies on coadministration of a mRNA-based COVID-19 and a seasonal quadrivalent influenza vaccine have reported a restriction of humoral Anti-SARS-CoV-2-Spike immune response in the coadministration group. These examinations were conducted with limited correspondence to real-life conditions and in smaller cohorts. Additionally, these former studies do not consider the important aspect of side effects as a possible direct effect of the prevention measure on the availability of public health care in combination with Anti-SARS-CoV-2-Spike IgG levels. In summary, the humoral immunogenicity and side effects of a coadministered third COVID-19 and a seasonal influenza vaccine are still unclear and the limited available data is not transferable to the general public.

*Added value of this study:* We performed the first large-scale real-life evaluation of humoral immunogenicity and side effects of COVID-19 and influenza vaccine coadministration in HCWs. Anti-SARS-CoV-2-Spike IgG levels were significantly lower in the coadministered cohort compared to the not coadministered control group, stratified by third COVID-19 vaccine (BNT162b2mRNA or mRNA-1273). Anti-SARS-CoV-2-Spike IgG post-vaccine elevation was lower among BNT162b2mRNA vaccinated HCWs than in those vaccinated with mRNA-1273 as a third COVID-19 vaccination. The influence of the seasonal quadrivalent influenza vaccine is evaluated in a cohort including 1,231 HCWs in total, covering a broad age range. Coadministration did not lead to an increase in side effects, which is a central requirement for considering the option of coadministration, given the role of HCWs as key personnel in maintaining health care capacities.

*Implications of all the available evidence:* Our data suggest, that coadministration of third mRNA-based COVID-19 and quadrivalent seasonal influenza vaccine is safe and immunogenic, although it leads to a slightly reduced Anti-SARS-CoV-2-Spike antibody formation. While the clinical impact of the observed reduction in humoral Anti-SARS-CoV-2-Spike immune response for protection against SARS-CoV-2 infection and severe COVID-19 disease is still unclear, influenza vaccination remains an important infection prevention measure, especially among highly exposed HCWs. The coadministration does not increase side effects but may improve vaccination rate. A higher-dosed mRNA-based COVID-19 vaccine may compensate for the restricted immunogenicity in case of seasonal influenza vaccine coadministration. Our results will support the development of public health recommendations for coadministration of COVID-19 and influence vaccines in anticipation of the imminent infection waves in the coming winter season.

## 1 Introduction

Influenza is one of the most common respiratory tract infections, especially in the northern hemisphere during the winter, and relevantly contributes to morbidity and mortality. ^1^ The COVID-19 pandemic adds as newly emerged viral respiratory seasonal infection with its severe burden of disease and underlines the need for prevention strategies. Due to the community-based COVID-19 mitigation measures, seasonal influenza prevalence has been observed as being reduced in times of the COVID-19 pandemic. ^2^ Given the current revocation of public strategies for SARS-CoV-2 infection prevention, a continued relevant COVID-19 prevalence and the absence of an accompanying decline in influenza prevalence could be expected for the next winter season. ^3^ Vaccines have been established as an effective infection prevention measure for both, COVID-19 and seasonal influenza. ^4,5^

Immunological capability is particularly crucial for healthcare workers (HCWs), being highly exposed to SARS-CoV-2 as well as influenza viruses and for being in contact with vulnerable patient groups in their daily profession. ^6,7^ COVID-19 and influenza vaccination are strongly recommended for HCWs as occupational infection prevention measures. ^4,8,9^ Occupation associated infections with both viral agents depict a relevant impact on staff shortages due to illness and consequently on the maintenance of healthcare capacities and need to be prevented.

In addition to the well-established annual seasonal influenza vaccination, a third COVID-19 vaccination dose has proven to be an important measure in maintaining a reliable SARS-CoV-2 immunity. ^10^ A coadministration of COVID-19 and seasonal influenza vaccine may offer a time and cost effective strategy to increase the uptake of both vaccines.

This study examines the antibody-mediated immunogenicity of coadministration of mRNA-based COVID-19 and quadrivalent seasonal influenza vaccine in HCWs, considering side effects as well as the impact of coadministration on Anti-SARS-CoV-2-Spike IgG concentrations.

## 2 Methods

### 2.1 Study setting

The data presented is part of the prospective CoVacSer cohort study investigating SARS-CoV-2 immunity, quality of life and ability to work in HCWs after COVID-19 vaccination and/or SARS-CoV-2 infection. The CoVacSer study enrolment required a complete fulfilment of the inclusion criteria: (i) age ≥ 18 years, (ii) written consent form, (iii) 14 days minimal interval after first polymerase chain reaction (PCR) derived confirmation of SARS-CoV-2 infection and/or at least one dose of COVID-19 vaccination independent of vaccination regime, (iv) employment in healthcare sector.

The German federal COVID-19 vaccination campaign started on the 27^th^ of December 2020 with consistent extension of vaccination capacities. On account of the initial shortage of vaccine, the vaccination followed a precedence based, phased plan in which HCWs were assigned to the highest priority level. ^11,12^

Following the recommendation of the German constant vaccination committee (STIKO), COVID-19 booster vaccinations were officially established at least six months after the second dose of COVID-19 vaccination since the 18^th^ of November 2021: for adults aged 18-30 years a single dose of BNT162b2mRNA (Comirnaty, BioNTech/Pfizer, Mainz/Germany, New York/USA; 30µg mRNA) was recommended, for individuals aged ≥30 years a single dose of BNT162b2mRNA or half a dose of mRNA-1273 (Spikevax, Moderna, Cambridge/USA; 50μg mRNA). Both heterogeneous as well as homologous vaccine combinations in the individual vaccination plans were recommended depending on availability of mRNA-based COVID-19 vaccines. ^12^

For this substudy, only participants who had received two doses of BNT162b2mRNA (30µg mRNA each) as basic SARS-CoV-2 immunisation without any SARS-CoV-2 infection convalescence were included. Either BNT162b2mRNA (30µg mRNA) or mRNA-1273 (50µg mRNA) was administered as third vaccination dose. Distribution of the two vaccines was based upon vaccine availability and the national recommendation to prefer BNT126b2mRNA administration as COVID-19 booster vaccine in individuals younger than 30 years.

HCWs who received a third COVID-19 vaccination could opt for a simultaneous influenza vaccination injected intramuscularly into the deltoid muscle of the opposite arm. The seasonal quadrivalent Influvac Tetra vaccine 2021/2022 (Abbott Biologicals, Olst/Netherlands) was used for influenza vaccination. Those HCWs who did not choose a coadministration of both vaccines, received either no seasonal influenza vaccination or an influenza vaccine at least 14 days apart from the COVID-19 booster vaccination.

### 2.2 Data collection

The data was collected between 1^st^ of October 2021 and 31^st^ of January 2022 over the course of the fourth and the beginning of the fifth wave of the COVID-19 pandemic in Germany. ^13^

Serum blood samples combined with the CoVacSer study questionnaire were collected between 14 and 90 days after the COVID-19 booster vaccination. The CoVacSer study survey queries socio-demographic aspects and individual risk factors, containing the World Health Organisation Quality of Life (WHOQOL-BREF) questionnaire as well as the Work Ability Index (WAI). ^14,15^ Predominantly HCWs from a single tertiary care hospital as well as some HCWs from surrounding hospitals and private practices were recruited consecutively.

Only blood samples with a signed written consent form and a completed linked questionnaire were considered. REDCap (Research Electronic Data Capture, projectredcap.org) was used as technological platform for the questionnaire recording. ^16^

In the context of pseudonymisation, blood samples were assigned to the study survey based on date of birth and dates of COVID-19 vaccination.

### 2.3 SARS-CoV-2 IgG ELISA

The measurement of Anti-SARS-CoV-2-Spike IgG levels was performed using the SERION ELISA *agile* SARS-CoV-2 IgG (SERION diagnostics, Wuerzburg, Germany) as an enzyme linked immunoassay. Due to previous performance evaluation studies, this serological assay was found to be reliable in terms of neutralisation activity compared with other available tests. ^17,18^

The photometric measurement of extinction values was carried out with Dynex Opsys MR(tm) Microplate Reader and Revelation Quick Link (Dynex technologies, Chantilly VA, USA) at 405 nm wavelength. Detected extinction values were converted to Serion IgG units per ml (U/ml) as producer specific units in use of the software easyANALYSE (SERION diagnostics). Consequently, the internationally established unit Binding Antibody Units per ml (BAU/ml) were calculated using the factor 2·1 in accordance with manufacturer’s information. ^19^

IgG levels beyond the threshold of 31·5 BAU/ml report at least a moderate neutralisation capacity. ^17^ For the measurement of Anti-SARS-CoV-2-Spike-IgG levels beyond the maximum limit of 250·0 U/ml (>525·0 BAU/mL), a dilution series containing dilution factors both 10 and 100 was conducted. Thus, the measurable spectrum of SERION ELISA *agile* SARS-CoV-2 IgG could be broadened.

### 2.4 Ethical approval

The study protocol was approved by the Ethics committee of the University of Wuerzburg in accordance with the Declaration of Helsinki (file no. 79/21).

### 2.5 Statistics

The statistical analyses were performed with the statistical programming language R (version 3.1.2). ^20^

Anti-SARS-CoV-2-Spike IgG concentrations were logarithmised and statistical analyses were performed on the logarithmised concentrations (*Supplementary Figure S2*). Based on a multiple linear regression analysis, differences in Anti-SARS-CoV-2-Spike levels were calculated. Besides coadministration, additional factors which might be associated with Anti-SARS-CoV-2-Spike IgG levels were incorporated into the analysis. Thus, information about age, gender, cigarette smoking, side effects after third COVID-19 vaccine administration, time between vaccination and serum sampling, being a high-risk subject (self-reported, according to participants’ self-assessment), and the administered vaccine (BNT162b2mRNA or mRNA-1273) were considered.

The parameters of the multiple linear regression model were estimated using a generalised least square fit, performed with the R package *nlme*. ^21,22^ Starting from the regression model, statistical differences between the subgroups of the independent categorical variables were calculated with the estimated marginal means. Using the Tukey test statistics, pairwise post hoc tests were performed to expose statistically significant differences. The *emmeans*^23^ package was used to estimate marginal means and to calculate pairwise differences.

Investigating the role of side effects due to coadministered vaccinations, pairwise Fisher exact tests were performed on the number of participants reporting side effects. The reported side effects were defined within five different groups: local reactions, headaches, muscle pain, fever and/or chills, fatigue and other side effects.

To correct against multiple testing, p-values were adjusted using the Benjamini-Yekutielie procedure. ^24^ Only adjusted p-values below a significance level of 0·05 were considered statistically significant.

## 3 Results

### 3.1 Study population

Between 1^st^ of October 2021 and 31^st^ of January 2022, 1,231 participants of the CoVacSer study received a third dose of mRNA-based COVID-19 vaccine (BNT162b2mRNA or mRNA-1273) as a booster (*Supplementary Figure S1*).

20·2% (249/1,231) of all participants were simultaneously coadministered with a third dose of COVID-19 vaccine and a seasonal quadrivalent influenza vaccine dose. Among those 249 participants who opted for coadminstration of the COVID-19 and influenza vaccines, 90·4% (225/249) received BNT162b2mRNA, 9·6% (24/225) mRNA-1273.

79·8% (982/1,231) of all participants were vaccinated with a mRNA-based COVID-19 vaccine only, forming the no coadministration control cohort without concurrent seasonal influenza vaccination. Within this control group, 78·5% (771/982) received BNT162b2mRNA and 21·5% (211/982) mRNA-1273 as COVID-19 booster vaccine (*Figure 1, Supplementary Figure S3*).

**Figure 1:**
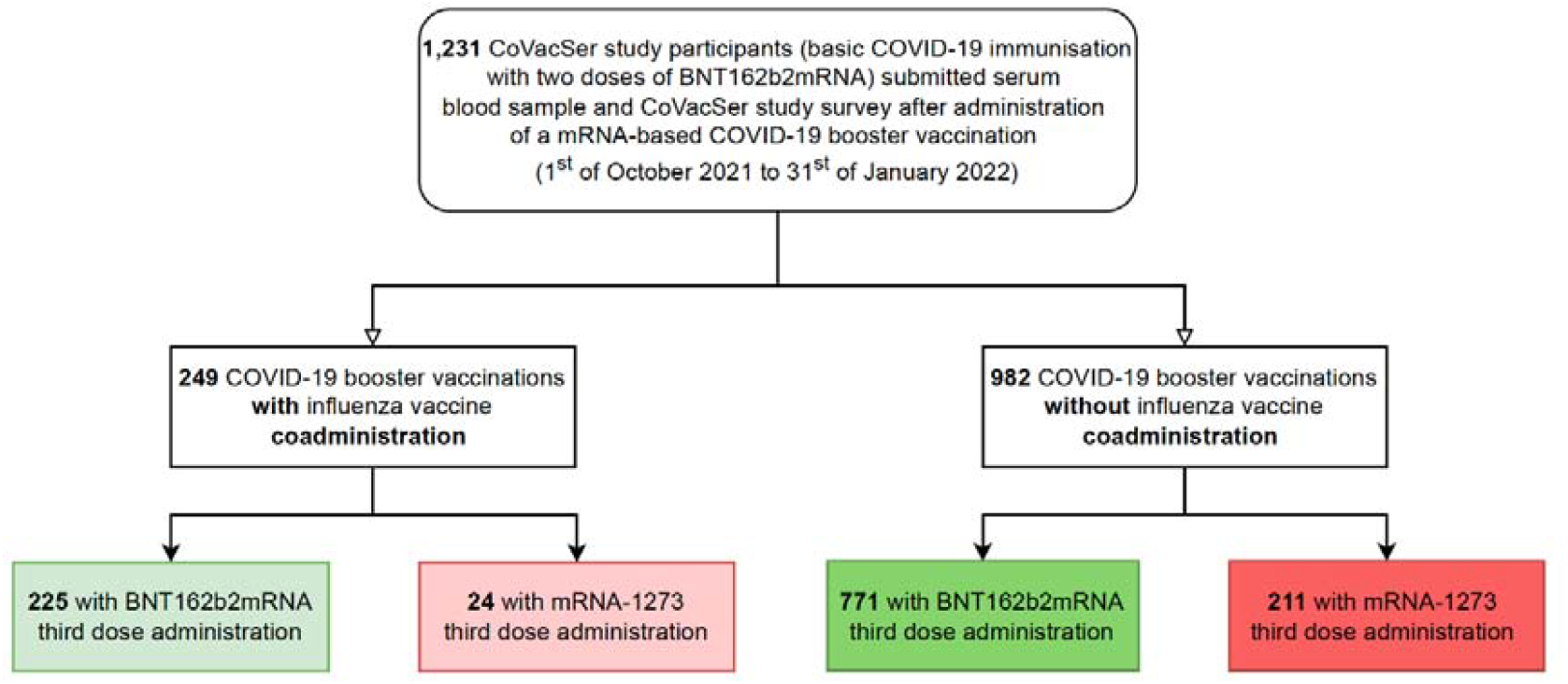
Enrolment of study participants and their allocation to subgroups concerning coadministration of COVID-19 booster and seasonal influenza vaccination.

A detailed comparative characterisation of the two groups with and without coadministration of seasonal influenza vaccine is shown in *Table 1*.

**Table 1:** Comparative characterisation of study groups with and without influenza vaccine coadministration. Comparisons of continuous features were performed with the non-parametric Wilcoxon test, while comparisons of gender proportions were performed by Fisher exact test. All p values were corrected against multiple testing.

| Characteristic | Coadministration | No Coadministration | Odds ratio | p |
| --- | --- | --- | --- | --- |
| Number of included respondents | 249 (20.2%) | 982 (79.8%) | - |  |
| Median age [years] (± IQR) | 38 (30-51) | 41 (31-53) | - | 0.40 |
| Weight [kilogram] (± IQR) | 70 (62-83) | 68 (60-80) | - | 0.32 |
| Height [centimetres] (± IQR) | 170 (165-176) | 168 (164-174) | - | 0.13 |
| BMI [kilogram/m<sup>2</sup>] (± IQR) | 24 (22-27) | 24 (21-27) | - | 0.05 |
| *Number of female* | 187 (75.1%) | 828 (84.3%) | 0.56 | < 0.01 |
| *Number of male* | 62 (24.9%) | 154 (15.7%) |  |  |
| *Interval between first and second COVID-19 vaccine administration [days] (± IQR)* | 21 (21-21) | 21 (21-21) | - | 1.00 |
| *Interval between second and third COVID-19 vaccine administration [days] (± IQR)* | 260 (231-288) | 267 (243-291) | - | 0.13 |
| *Interval between third COVID-19 vaccine administration and study participation [days] (± IQR)* | 18 (15-24) | 18 (15-24) | - | 1.00 |

### 3.2 Influence of COVID-19 booster and influenza vaccine coadministration on Anti-SARS-CoV-2-Spike IgG levels

The Anti-SARS-CoV-2-Spike IgG levels ranged from 239·1 to 18,541·5 BAU/ml (median 1,605·0 BAU/ml, IQR: 1,078·0-2,504·7 BAU/ml) in the group of COVID-19 and influenza coadministered study participants. In the cohort vaccinated exclusively with a COVID-19 booster dose, Anti-SARS-CoV-2-Spike IgG levels from 17·7 to 21,287·9 BAU/ml were determined (median: 2,150·2 BAU/ml, IQR 1,341·1-3,242·3 BAU/ml).

Median Anti-SARS-CoV-2-Spike IgG levels were 34·0% higher in HCWs who were not coadministered with the influenza vaccine simultaneously to the COVID-19 vaccination (p<0·01), *Figure 2*).

**Figure 2:**
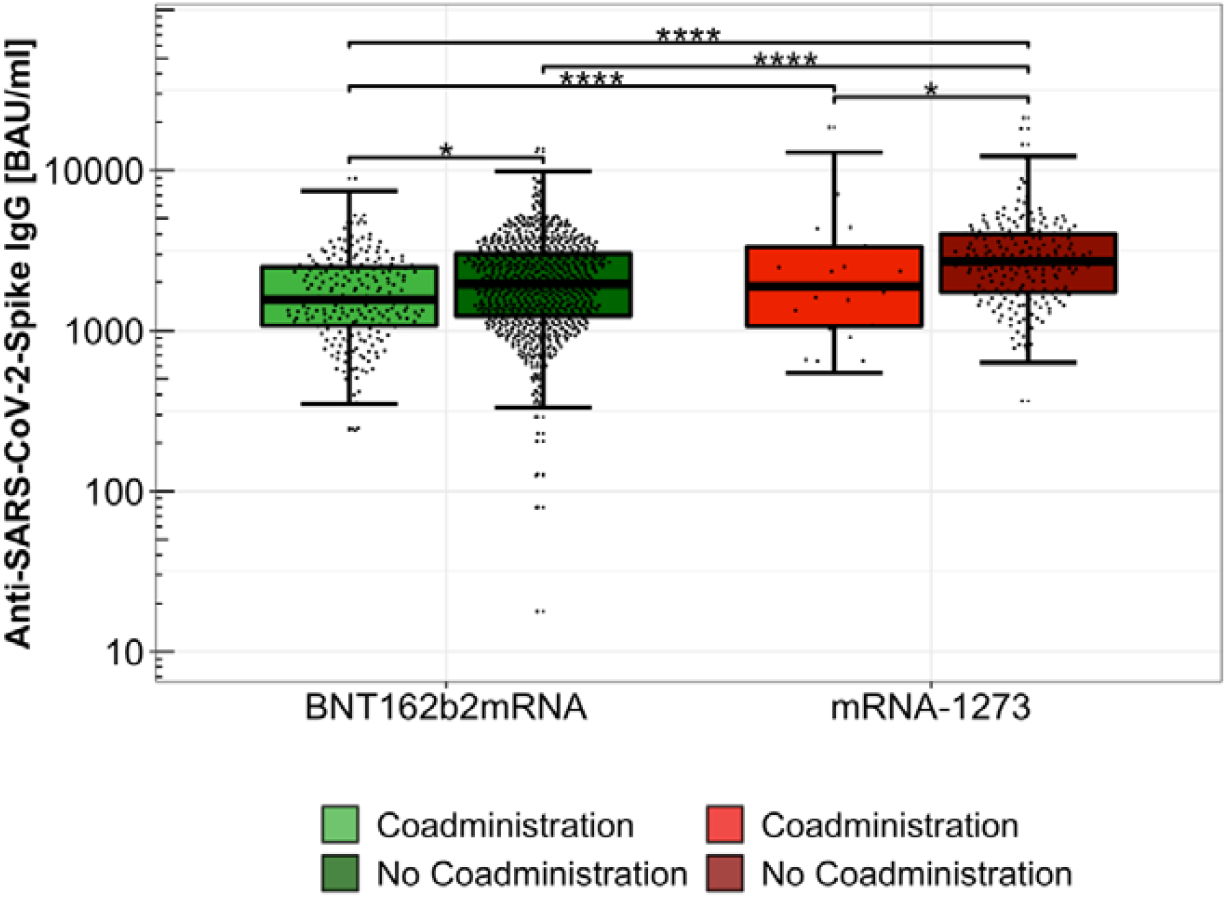
Anti-SARS-CoV-2-Spike IgG concentrations after third dose of COVID-19 vaccine with or without influenza vaccine coadministration, separated for COVID-19 booster vaccine (BNT162b2mRNA or mRNA-1273), logarithmically scaled. ****: p < 0·0001 *: p < 0·05 BAU/ml: Binding Antibody Units per millilitre

Subgroup analysis showed that in case of Influvac Tetra and BNT162b2mRNA coadministration, the median Anti-SARS-CoV-2-Spike IgG was 1,560·7 BAU/mL (IQR: 1,078·0-2,493·6 BAU/ml), compared to 1,955·1 in participants only administered with BNT162b2mRNA (IQR: 1,234·6-3,022·9 BAU/ml). For coadministration of Influvac Tetra and mRNA-1273, a median of 1,891·1 BAU/mL (IQR: 1,068·2-3,324·0 BAU/ml) could be observed, whereas in mRNA-1273-only vaccinated respondents, the median Anti-SARS-CoV-2-Spike IgG level obtained was 2,709·8 BAU/ml (IQR: 1,735·4-4,001·2 BAU/ml). Anti-SARS-CoV-2-Spike IgG concentrations were significantly higher without influenza vaccine coadministration, for both BNT162b2mRNA (p<0·05) as well as mRNA-1273 (p<0·05) administered participants.

Independent of influenza vaccine coadministration, mRNA-1273 induced significantly higher Anti-SARS-CoV-2-Spike IgG concentrations than BNT162b2mRNA (p<0·0001; *Figure 2*; *Supplementary Figure S6*).

### 3.3 Effects of COVID-19 booster and influenza vaccine coadministration on vaccine related side effects

Overall, in the sub-cohort of study participants coadministered with COVID-19 and influenza vaccines, all listed adverse events were reported less frequently than in participants without coadministration.

Among the coadministered participants, 81·1% (202/249) reported vaccine-related side effects compared to 87·3% (857/982) in the solely COVID-19 booster vaccinated group. After vaccine administration, 59·8% (149/249) of coadministered participants suffered from local reactions, 37·3% (93/249) from headache, 34·1% (85/249) from muscle pain, 26·1% (65/249) from fever and/or chills, 47·8% (119/249) from fatigue and 12·9% (32/249) from other complaints not covered in the previously formed categories in the questionnaire.

Among solely COVID-19 booster vaccinated study participants, 62·0% (609/982) reported local reactions, 43·3% (425/982) headache, 41·8% (410/982) muscle pain, 29·5% (290/982) fever and/or chills, 52·6% (517/982) fatigue and 15·1% (148/982) other complaints after vaccination. Numerically, all queried side effects were less common in the coadministration group but for none of the side effects this difference was statistically significant (p>0·05; *Figure 3*).

**Figure 3:**
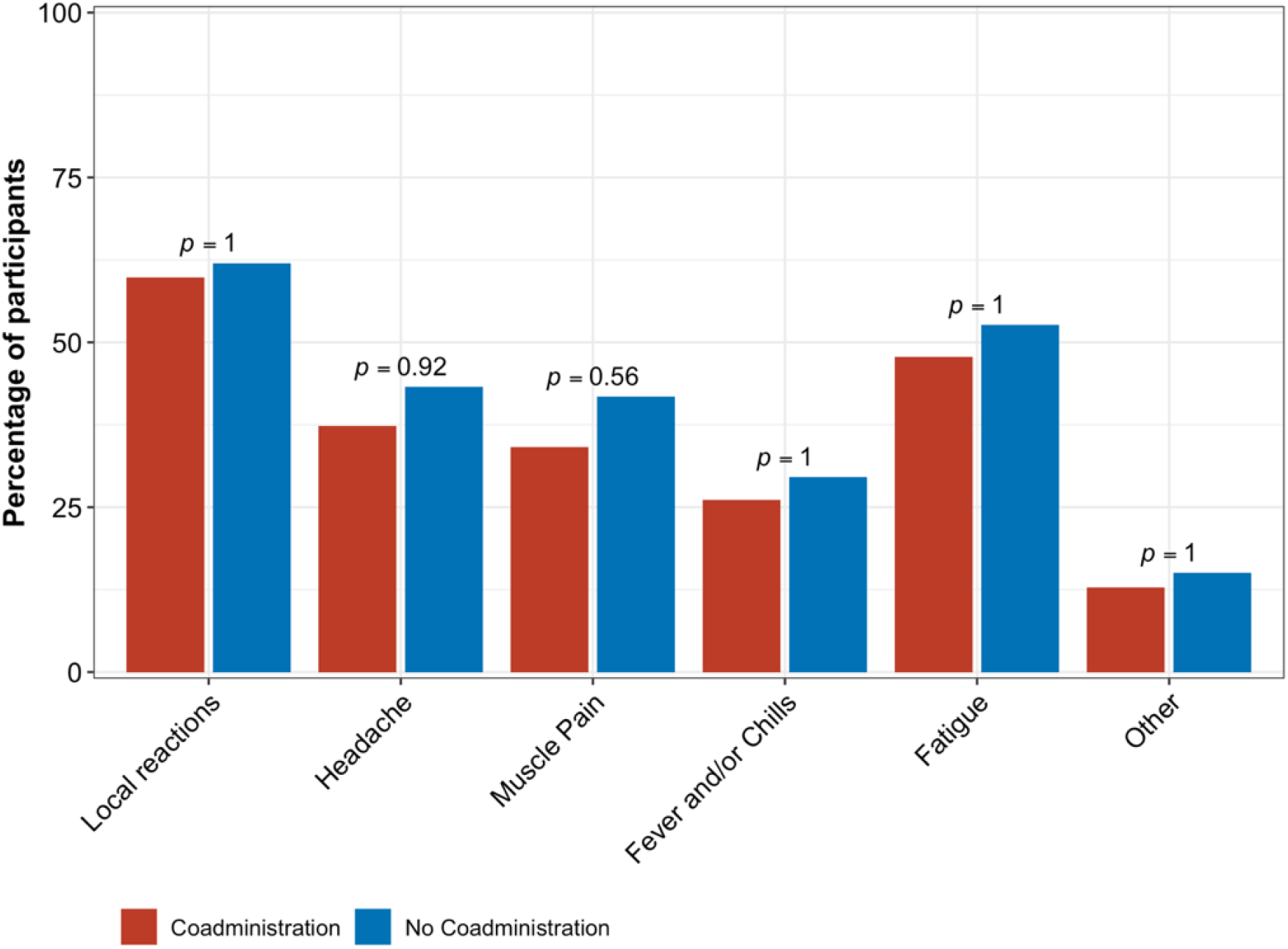
Frequency of vaccine-related side effects in study participants after a third dose of COVID-19 vaccine with (red bars) or without (blue bars) concurrent administration of a seasonal quadrivalent influenza vaccine.

## 4 Discussion

A third dose of COVID-19 vaccine (i.e. booster vaccination) induced high Anti-SARS-CoV-2-Spike IgG levels, also when a seasonal quadrivalent influenza vaccine was coadministered. The coadministration of a COVID-19 booster vaccination and a seasonal influenza vaccine did not lead to increased reporting of vaccine-related side effects.

However, median Anti-SARS-CoV-2-Spike IgG levels were by more than 25% lower in individuals who had received their third mRNA-based COVID-19 vaccination simultaneously with a seasonal quadrivalent influenza vaccine compared to solely COVID-19 booster vaccinated individuals in the control cohort. This fact was observed for both mRNA-based vaccines, BNT162b2mRNA and mRNA-1273. Consequently, coadministration was associated with a decreased humoral immune response to COVID-19 vaccination using mRNA vaccines, possibly due to parallel stimulation of the immune system by both vaccines. These findings are in line with decreased Anti-SARS-CoV-2 antibody titres observed in individuals receiving a combined protein subunit COVID-19 and influenza vaccine compared to individuals receiving a protein subunit COVID-19 vaccine without an influenza component.^25^ The clinical consequences of the impaired humoral immune response in case of coadministration in this high Anti-SARS-CoV-2-Spike IgG level range are still unclear, as it remains to be understood which Anti-SARS-CoV-2-Spike IgG levels are required for reliable protection against SARS-CoV-2 infection and severe COVID-19 disease. ^26^

Possibly, an adapted and augmented COVID-19 vaccine dosage in case of influenza vaccine coadministration might compensate for the lower levels of Anti-SARS-CoV-2-Spike IgG after coadministration of both vaccines. The comparison of participants administered with the higher-dosed mRNA-1273 (50µg) COVID-19 booster vaccine compared to BNT162b2mRNA (30µg) confirms this assumption, by displaying similar Anti-SARS-CoV-2-Spike IgG levels after booster vaccination in the subgroup vaccinated with BNT162b2mRNA only and the subgroup coadministered with the influenza vaccine and mRNA-1273. Consequently, the higher-dosed mRNA-1273 booster vaccine might compensate for the limited increase in Anti-SARS-CoV-2-Spike IgG induced after coadministration of COVID-19 booster and influenza vaccine. However, a possible increase in adverse events should be carefully monitored and needs to be taken into consideration.

The limited increase of Anti-SARS-CoV-2-Spike IgG concentrations after COVID-19 booster vaccination among coadministered participants may explain the absence of increased side effects despite the enhanced immunisation stimulus from both viral agents. Potentially, those HCWs who had fewer side effects from the initial two vaccination doses were more willing to receive the influenza vaccination in addition to the COVID-19 booster vaccination. COVID-19 and influenza vaccine coadministration did not induce an increase in adverse events compared to the corresponding control of only COVID-19 booster vaccinated HCWs. This observation is in concordance with previous studies. ^25,27,28^ Coadministration does not increase side effects which is a meaningful reasoning for considering the option of coadministration, given the role of HCWs as key personnel in maintaining healthcare capacities.

The findings of this study should be considered in the light of the following possible limitations: Firstly, presented data was collected as non-randomised clinical trial, included study participants were not assigned by chance to the compared groups and opted based on their individual, subjective decision for coadministration. The reasons for the decisions were not evaluated. The study population was predominantly female (82·5%). This unequal gender distribution of enrolled participants reflects the current gender distribution of the workforce in the German public healthcare sector (75·5% female). ^29^ The share of male participants was slightly higher in the coadministration group compared to the control group. However, the multiple regression analysis modelling gender effects reveals that the group differences in Anti-SARS-CoV-2-Spike IgG concentrations cannot be explained by the gender distribution in the two groups of the participants. Additionally, this model did not detect any significant gender-specific differences in Anti-SARS-CoV-2-Spike IgG levels. The data presented enables a real-life analysis of COVID-19 and influenza vaccine coadministration, including the important aspect of side effects with its direct impact on public health care capacity. It should further be noted that vaccine-related side effects were assessed using self-report questionnaires. In the study presented, only the effects of coadministration on Anti-SARS-CoV-2-Spike IgG levels were examined, whereas the influenza-specific humoral immune response was not obtained.

Due to vaccine availability during the data collection period and participants’ individual reasons, COVID-19 booster vaccinations were administered with either BNT162b2mRNA or mRNA-1273 representing a point-of-care scenario. The influence on Anti-SARS-CoV-2-Spike IgG levels as well as vaccine-related side effects was evaluated separately for both vaccines. Thereby, we were able to provide relevant information for both mRNA-based COVID-19 vaccines, mRNA-1273 as well as BNT162b2mRNA, coadministered with seasonal influenza vaccine. Based on the limited local vaccine offer, respondents beyond the age of 60 years were not vaccinated with a high-dose influenza vaccine as recommended by the German constant vaccination committee (STIKO). ^30^ Therefore, possible consequences of coadministration of a high-dose influenza vaccine, including potential effects on humoral immune response and side effects, should be further investigated to improve the transferability of the results to the general population, especially to people of advanced age.

## 5 Conclusion

Significantly lower Anti-SARS-CoV-2-Spike IgG concentrations were obtained after coadministration of COVID-19 booster and seasonal quadrivalent influenza vaccine compared to COVID-19 booster vaccination without coadministered influenza vaccine. The potential clinical impact of a reduced humoral immune response against SARS-CoV-2 remains unknown, highlighting the urgent need for further investigation, especially in the light of the upcoming influenza season. ^2,31^

In the context of seasonal influenza vaccination campaigns, an important conclusion of our study is that coadministration of COVID-19 and influenza vaccination does not increase side effects. This suggests that coadministration of COVID-19 and influenza vaccines is safe and will not jeopardise public healthcare capacities due to increased sick leave. In addition, the possibility of coadministration could increase the individual compliance and COVID-19 and influenza vaccination rates in the group of highly exposed HCWs.

## Data Availability

Additional data that underlie the results reported in this article, after de-identification (text, tables, figures, and appendices) as well as the study protocol, statistical analysis plan, and analytic code is made available to researchers who provide a methodologically sound proposal to achieve aims in the approved proposal on request to the corresponding author.

## Abbreviations

**PCR, HCWs**

## 6 Author contributions

All authors had unlimited access to all data. Isabell Wagenhäuser, Julia Reusch, Alexander Gabel, Nils Petri and Manuel Krone take responsibility for the integrity of the data and the accuracy of the data analysis.

Conception and design: Lâm, Almanzar, Prelog, Frey, Schubert-Unkmeir, Dölken, Frantz, Kurzai, Vogel, Petri, Krone M.

Anti-SARS-CoV-2-Spike IgG concentration determination: Wagenhäuser, Reusch.

Trial management: Wagenhäuser, Reusch, Höhn, Petri, Krone M.

Statistical analysis: Wagenhäuser, Reusch, Gabel, Krone L.B., Petri, Krone M.

Obtained funding: Kurzai, Vogel.

First draft of the manuscript: Wagenhäuser, Reusch, Gabel, Petri, Krone M.

Reviewing and modifying the manuscript and approving its final version: Höhn, Lâm, Almanzar, Prelog, Krone L.B., Frey, Schubert-Unkmeir, Dölken, Frantz, Kurzai, Vogel.

## 10 Acknowledgements

We thank the technical assistants of the serological diagnostic laboratory for sharing their laboratory, and especially for their help and advice.

## Supplementary Material

**Supplementary Figure S1:**
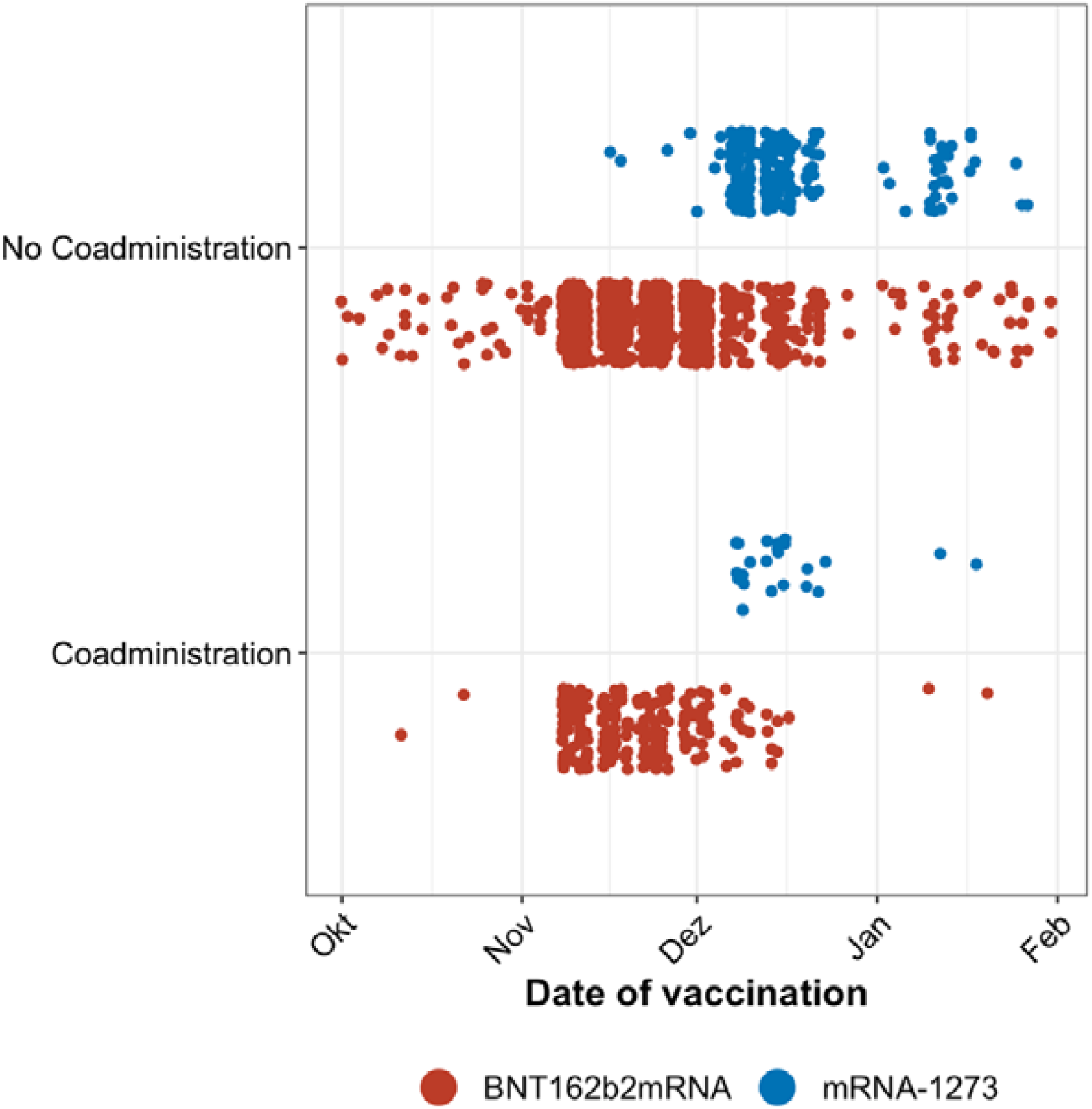
Temporal distribution of COVID-19 booster vaccination, COVID-19 vaccine separated. Temporal display of COVID-19 booster vaccination date visualised as point distribution, separated by COVID-19 vaccines (red dots representing BNT162b2mRNA, blue dots mRNA-1273).

**Supplementary Figure S2:**
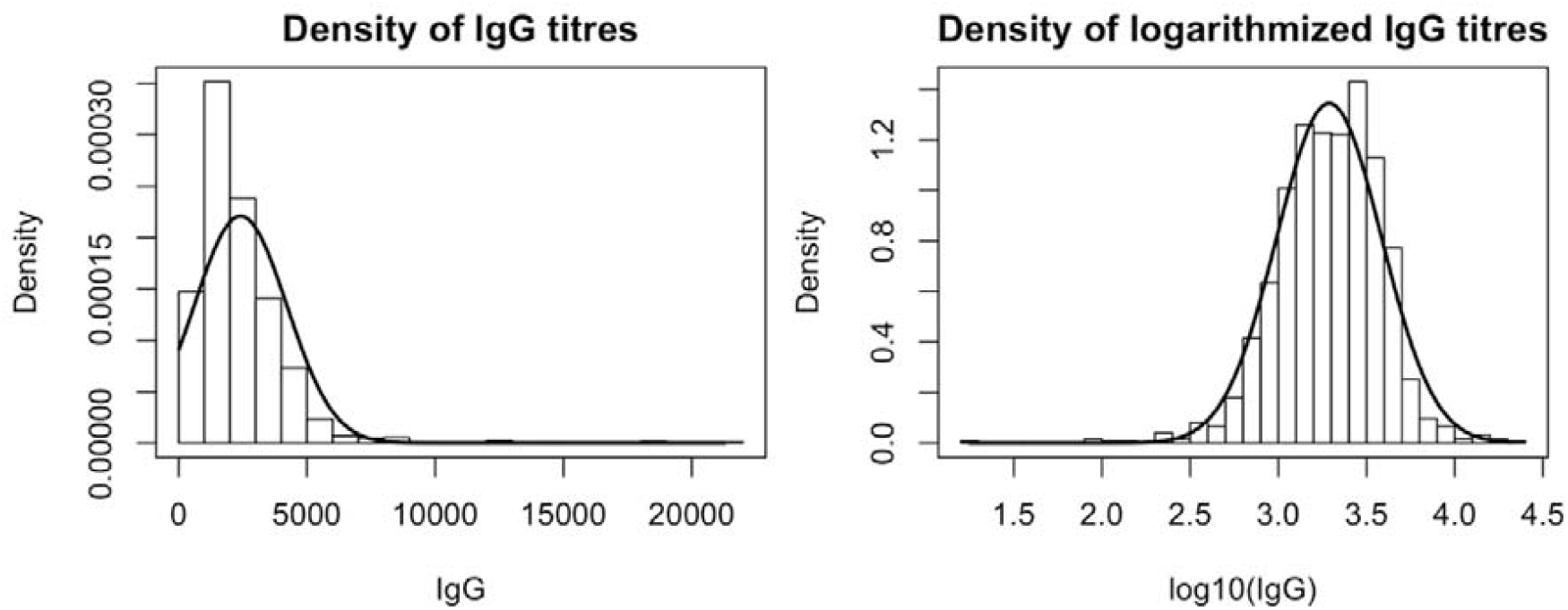
Distribution of Anti-SARS-CoV-2-Spike IgG levels. Density histogram of Anti-SARS-CoV-2-Spike IgG concentrations and of logarithmically scaled Anti-SARS-CoV-2-Spike IgG concentrations. Obtained logarithmised IgG levels of included study participants are nearly normal distributed.

**Supplementary Figure S3:**
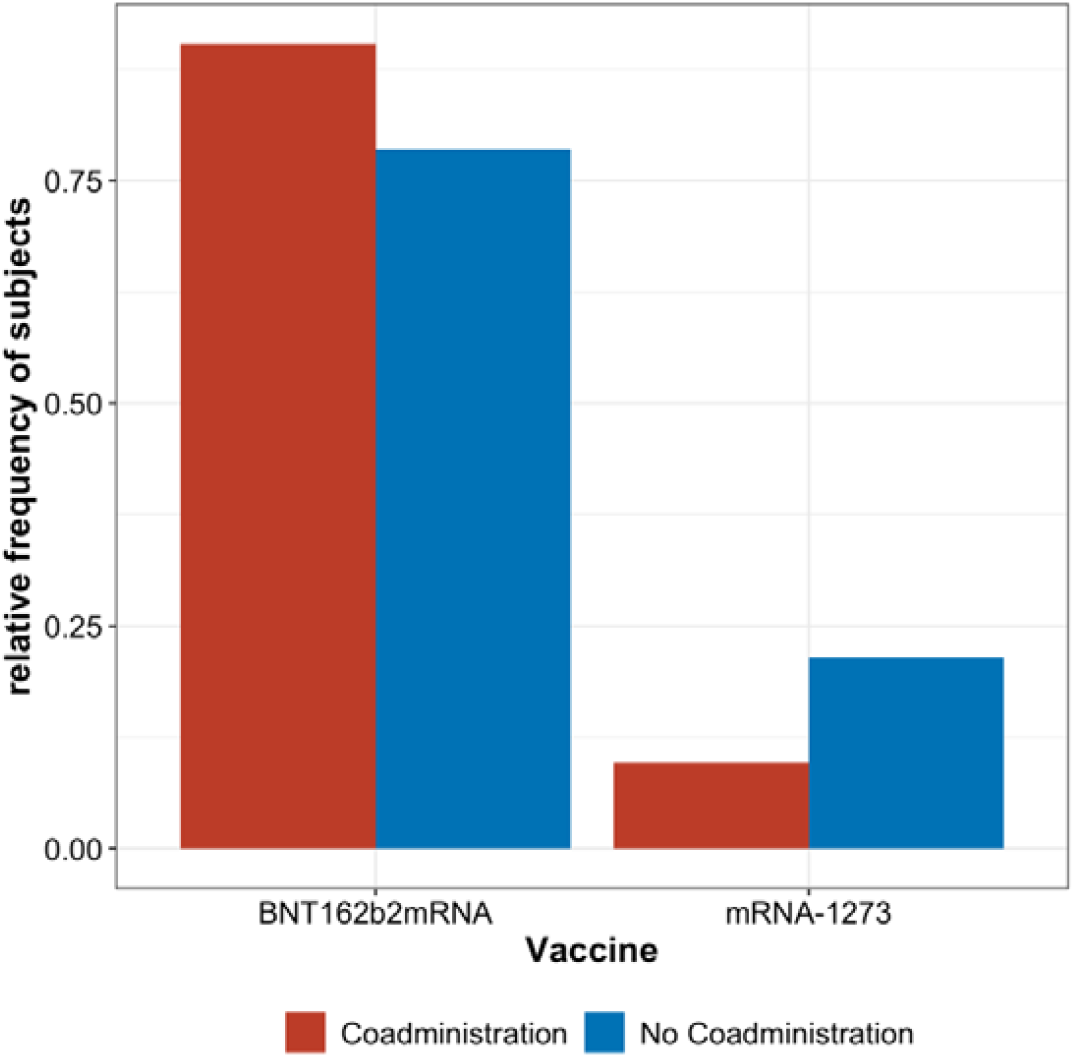
Relative frequency of COVID-19 vaccines within coadministrated and not coadministrated HCWs. Relative frequencies of vaccinated HCWs separated by COVID-19 booster vaccines. Frequencies were normalized over the groups of participants with (red) and without influenza coadministration (blue).

**Supplementary Figure S4:**
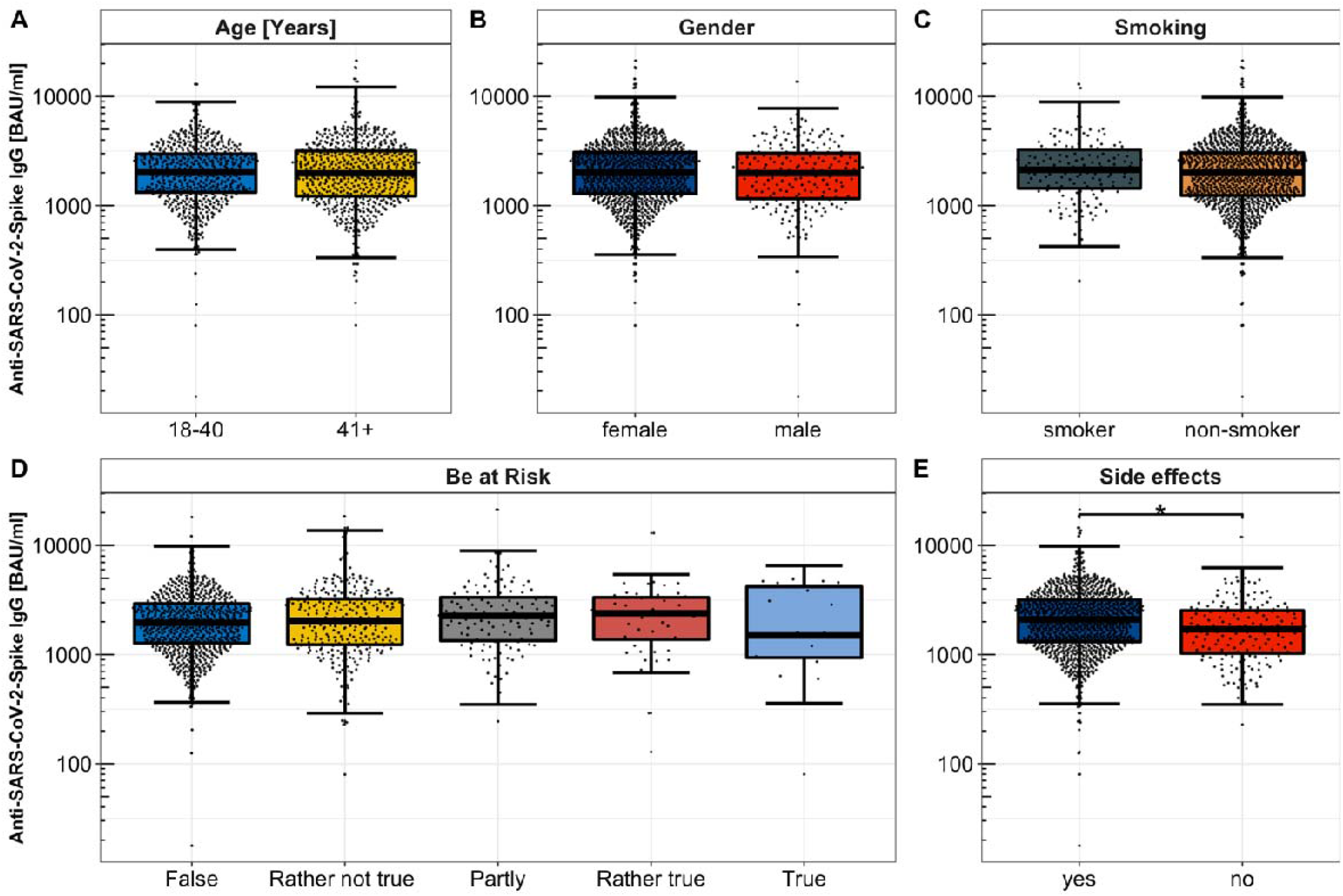
Comparison of Anti-SARS-CoV-2-Spike IgG concentrations between groups of remaining factors. Pairwise comparisons of Anti-SARS-CoV-2-Spike IgG concentrations between subgroups of remaining factors. *: p < 0·05 BAU/ml: Binding Antibody Units per millilitre

**Supplementary Figure S5:**
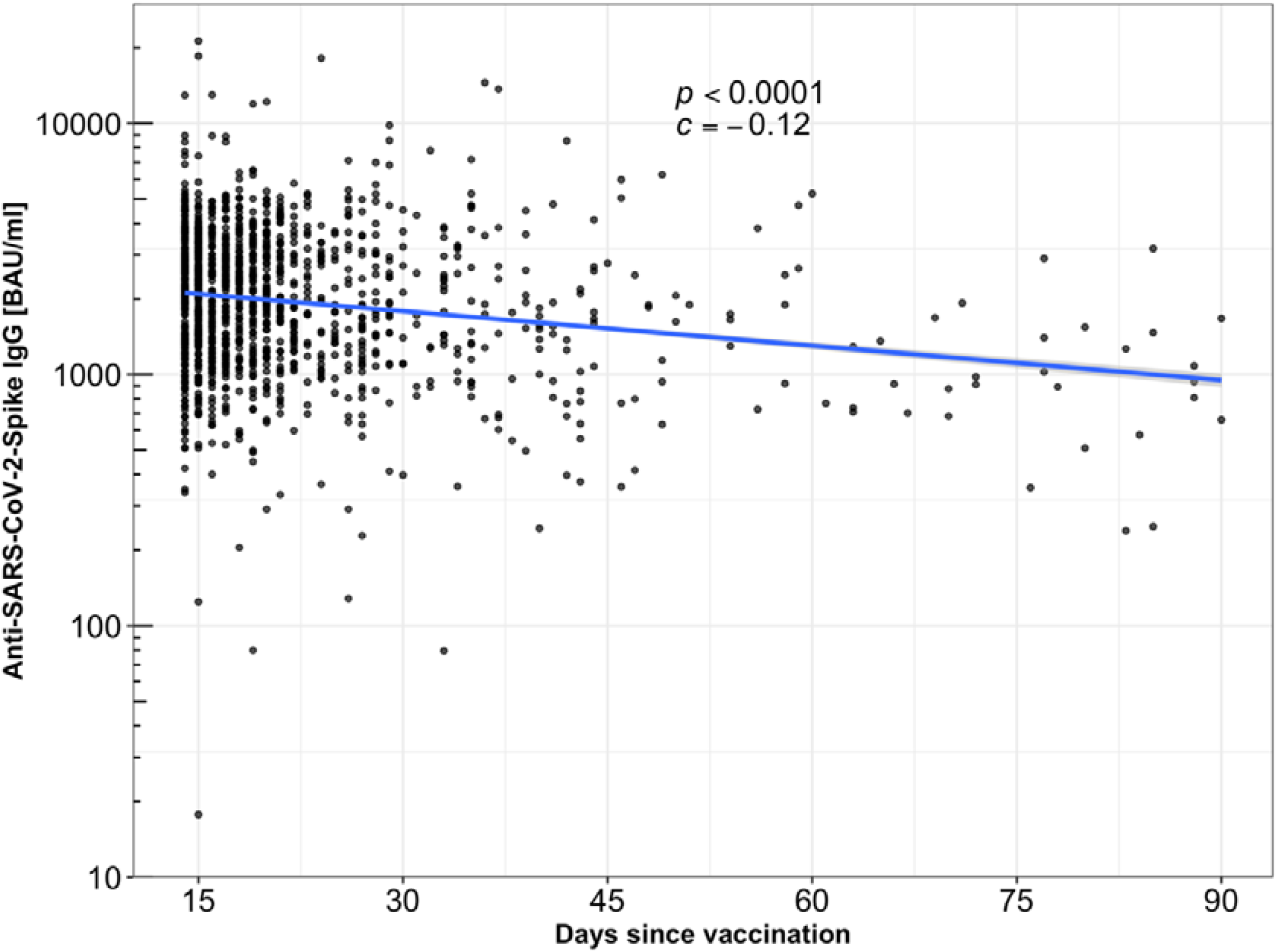
Temporal decline of Anti-SARS-CoV-2-Spike IgG after COVID-19 booster vaccination. Time course of Anti-SARS-CoV-2-Spike IgG levels as a function of days since third dose of COVID-19 vaccination. BAU/ml: Binding Antibody Units per millilitre

**Supplementary Figure S6:**
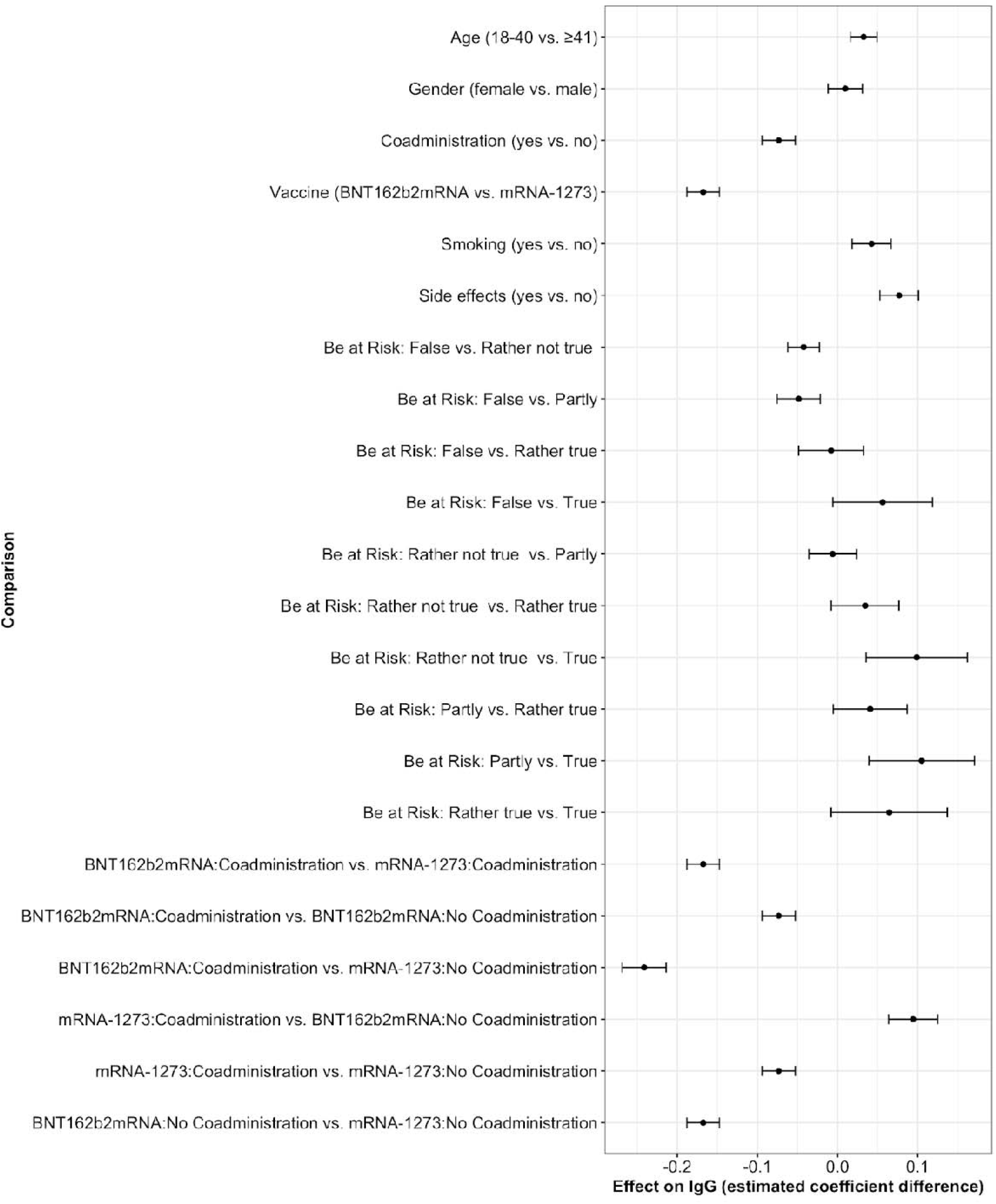
Pairwise comparison of Anti-SARS-CoV-2-Spike IgG influencing factors. Results of pairwise comparisons of Anti-SARS-CoV-2-Spike IgG influencing factors. The abscissa shows the differences of the estimated marginal means while the ordinate shows all pairwise comparisons with statistically significant differences. The points represent the differences of the estimated marginal means, while the whiskers represent the estimated standard errors.

**Supplementary Table S7:** Pairwise comparison of Anti-SARS-CoV-2-Spike IgG influencing factors.

|  | Groups |  | IgG Median (IQR) |  | p - value |  |
| --- | --- | --- | --- | --- | --- | --- |
| *Factor* | Group 1 | Group 2 | Group1 | Group2 | uncorrected | adjusted |
| **Age** | 18-40 | 40 | 2052.6 (1321.9-2979.5) | 1976.4 (1221.5-3172.6) | 0.05 | 0.41 |
| **Gender** | female | male | 2024.4 (1287.3-3082.1) | 1980.8 (1157.2-3029.7) | 0.66 | 1.00 |
| **Coadministration** | yes | no | 1605 (1078-2504.7) | 2150.2 (1341.1-3242.3) | < 0.001 | < 0.01 |
| **COVID-19 vaccine** | BNT162b2mRNA | mRNA-1273 | 1888.4 (1192.6-2916.8) | 2655.8 (1698.7-3987.9) | < 0.0001 | < 0.0001 |
| **Smoking** | yes | no | 2134 (1437.2-3264.5) | 1997.1 (1237.3-3051.2) | 0.08 | 0.6 |
| **Side effects** | yes | no | 2103.8 (1310.8-3184.6) | 1702.7 (1027.9-2534.7) | < 0.01 | 0.02 |
| **Be at risk:** |  |  |  |  |  |  |
|  | false | rather false | 1955.1 (1270.5-2935.4) | 2037.6 (1232.6-3238.8) | 0.20 | 1.00 |
|  | false | partly | 1955.1 (1270.5-2935.4) | 2258 (1339.7-3343.7) | 0.37 | 1.00 |
|  | false | rather true | 1955.1 (1270.5-2935.4) | 2396.9 (1377-3341.1) | 1.00 | 1.00 |
|  | false | true | 1955.1 (1270.5-2935.4) | 1493.7 (936.2-4202.1) | 0.90 | 1.00 |
|  | rather false | partly | 2037·6 (1232·6-3238·8) | 2258 (1339·7-3343·7) | 1·00 | 1·00 |
|  | rather false | rather true | 2037·6 (1232·6-3238·8) | 2396·9 (1377-3341·1) | 0·93 | 1·00 |
|  | rather false | true | 2037·6 (1232·6-3238·8) | 1493·7 (936·2-4202·1) | 0·52 | 1·00 |
|  | partly | rather true | 2258 (1339·7-3343·7) | 2396·9 (1377-3341·1) | 0·90 | 1·00 |
|  | partly | true | 2258 (1339·7-3343·7) | 1493·7 (936·2-4202·1) | 0·50 | 1·00 |
|  | rather true | true | 2396·9 (1377-3341·1) | 1493·7 (936·2-4202·1) | 0·90 | 1·00 |
| **Third dose COVID-19 vaccine and coadministration:** |  |  |  |  |  |  |
|  | BNT162b2mRNA<br>Coadministration | mRNA-1273<br>Coadministration· | 1560·7 (1078-2493·6) | 1891·1 (1068·2-3324) | < 0·0001 | < 0·0001 |
|  | BNT162b2mRNA<br>Coadministration · | BNT162b2mRNA<br>No Coadmin· | 1560·7 (1078-2493·6) | 1955·1 (1234·6-3022·9) | < 0·01 | 0·02 |
|  | BNT162b2mRNA<br>Coadministration · | mRNA-1273 No<br>Coadministration | 1560·7 (1078-2493·6) | 2709·8 (1735·4-4001·2) | < 0·0001 | < 0·0001 |
|  | mRNA-1273<br>Coadministration | BNT162b2mRNA<br>No<br>Coadministration | 1891·1 (1068·2-3324) | 1955·1 (1234·6-3022·9) | 0·01 | 0·11 |
|  | mRNA-1273<br>Coadministration | mRNA-1273 No<br>Coadministration | 1891·1 (1068·2-3324) | 2709·8 (1735·4-4001·2) | < 0·01 | 0·02 |
|  | BNT162b2mRNA<br>No Coadministration | mRNA-1273 No<br>Coadministration | 1955·1 (1234·6-3022·9) | 2709·8 (1735·4-4001·2) | < 0·0001 | < 0·0001 |

